# Serological prevalence of antibodies to SARS CoV-2 amongst cancer centre staff

**DOI:** 10.1101/2020.05.16.20099408

**Authors:** Karol Sikora, Ian Barwick, Ceri Hamilton

**Author notes:** Corresponding author’s.

## Abstract

**Objectives:** the aim of this study was to test Rutherford Health (RH) staff for the presence of SARS CoV-2 antibodies to reduce the risk of infection to cancer patients.

**Setting:** Between 14 and 24 April 2020 we tested 161 staff at four locations: our cancer centres in Reading - Berkshire, Newport - S Wales, Liverpool - Merseyside, and Bedlington in Northumberland.

**Participants:** Testing was available to all staff who were on site at the four locations named above at the time the study was carried out. 161 staff (80 men, 81 women) gave voluntary consent to have the tests and all testing gave rise to valid results.

**Interventions:** We used the South Korean test for antibodies to SARS CoV-2: Sugentech SGTi-flex COVID-19 IgM/IgG^1^. For each test, blood was collected and added to the sample well of the test cassette and buffer solution added. The test result was legible after 15 minutes. Outcome measures: The number of tests positive for the presence of antibodies was the primary outcome measure. The ratio of tests positive for the presence of IgM antibodies versus IgG antibodies was the secondary outcome measure.

**Results:** Between 14 and 24 April 2020, 161 staff (age m = 43) were tested at four Rutherford Cancer Care centres that offer proton beam therapy, radiotherapy and chemotherapy. Out of 161, 12 samples (7.50%) tested positive of which 7 samples (4.35%) detected IgM only, 2 samples (1.24%) detected IgG only and 3 samples (1.86%) detected both IgM and IgG.

**Conclusions:** The low seroconversion rate in the sample population limits the current utility of the test as a way of reducing risk to vulnerable patient populations but longitudinal retesting will provide further data.

Strengths and limitations of the study
- This is the first UK study on SARS CoV-2 antibody testing using the Sugentech SGTi-flex COVID-19 IgM/IgG in the workplace;
- This is the first UK study testing a population of cancer centre staff for SARS CoV-2 antibodies;
- This study builds on similar studies in other countries^3-4^, albeit with a smaller sample;
- Data on previous clinical symptoms is not included. We are in the process of revising consent paperwork for the test to include a question about previous symptoms like temperature, dry/persistent cough;
- Patient data is not included. We will consider testing patients once the pilot test with staff has reached conclusions about the efficacy and value of the test.

## Introduction

Previous work during the SARS outbreak in 2003^2^, reported that for the first 2-week period after the onset of infection, genetic testing based on reverse transcriptase polymerase chain reaction (RT-PCR) was the most sensitive method for the virus detection. However, in the convalescent phase, the detection of antibodies in the blood or serum specimens was more important. This is likely to be the case for SARS CoV-2.

Serological tests appear to hold the key for widespread population testing. These could determine the extent of the disease transmission through a population and possibly provide a route out of social distancing measures. Detecting the presence of antibodies to SARS-Cov-2 does not necessarily mean an individual would be immune from future infection. The market has been flooded by serological kits, many of them without the required specificity and sensitivity. These have been used in a haphazard manner and often by self-administration.

## Materials

At Rutherford Health, we began a pilot study to investigate whether antibodies could be detected in our staff at our four cancer centres providing day-care proton beam therapy, radiotherapy and chemotherapy. The aim was to see if we could reduce the risk of infection to patients from our frontline clinical team. We selected a South Korean test kit (Sugentech SGTi-flex COVID-19 IgM/IgG) based on the validation data supplied by both the manufacturer and from experience in Germany. We also compared the results with a chemiluminescent immunoassay at a North-west London laboratory, which demonstrated excellent concordance with the kit assays.

## Patient and public involvement

No Rutherford Cancer Centre patient and no member of the public was involved in this study. All cancer centre staff were eligible to take part in the study on a voluntary basis. The clinical governance group of Rutherford Cancer CentresHealth waived the need for ethical approval as only fully consenting staff were involved in this project.

## Method

Over the period 14-24 April 2020, we tested 161 staff at our centres (average age of 43 years) using this assay and found 12 (7.50%) samples being identified as positive of which 7 samples (4.35%) detected IgM only, 2 samples (1.24%) detected IgG only and 3 samples (1.86%) detected both IgM and IgG. We compared the prevalence of antibodies amongst our staff to two recent studies. The first was from Santa Clara County^3^, California, USA where 3,300 people were tested with a 1.5% positive test rate and the second from Gangelt^4^, Germany where 1,000 people were tested with a 14% positive rate. This town had a very high prevalence of viral infection.

## Discussion

The hope was that these tests could be used by occupational health services to allow antibody positive workers to return to work and potentially for international travellers to avoid quarantine on arrival. However, the low seroconversion rate almost certainly precludes this. We will investigate this further by gathering longitudinal data from our staff over future weeks. Other immune responses could well be providing important lines of defence against this virus.

K. Sikora, I. Barwick and C. Hamilton contributed to the design and implementation of the research, to the analysis of the results and to the writing of the manuscript.

We declare no competing interests.

This study received corporate funding from Rutherford Health PLC for the purchase of the Sugentech SGTi-flex COVID-19 IgM/IgG testing kits and their transport from South Korea. No other additional funding was required to carry out the study.

Raw data can be accessed by emailing

## Data Availability

Raw data can be accessed by emailing

